# Dissecting clinical heterogeneity of bipolar disorder using multiple polygenic risk scores

**DOI:** 10.1101/2020.03.31.20044800

**Authors:** Brandon J. Coombes, Matej Markota, J. John Mann, Colin Colby, Eli Stahl, Ardesheer Talati, Jyotishman Pathak, Myrna M. Weissman, Susan L. McElroy, Mark A. Frye, Joanna M. Biernacka

**Affiliations:** Department of Health Sciences Research, Mayo Clinic, Rochester, MN; Department of Psychiatry and Psychology, Mayo Clinic, Rochester, MN; Department of Psychiatry, Columbia University Vagelos College of Physicians & Surgeons, New York, NY; Division of Molecular Imaging and Neuropathology, New York State Psychiatric Institute, New York, NY; Pamela Sklar Division of Psychiatric Genomics, Icahn School of Medicine at Mount Sinai, New York, NY; Medical and Population Genomics, Broad Institute, Cambridge, MA; Divisions of Translational Epidemiology, New York State Psychiatric Institute, New York, NY; Department of Healthcare Policy & Research, Weill Medical College, Cornell University, New York, NY; Department of Epidemiology, Mailman School of Public Health, Columbia University, New York, NY; Lindner Center of HOPE/University of Cincinnati, Cincinnati, OH

## Abstract

Bipolar disorder (BD) has high clinical heterogeneity, frequent psychiatric comorbidities, and elevated suicide risk. To determine genetic differences between common clinical sub-phenotypes of BD, we performed a systematic PRS analysis using multiple polygenic risk scores (PRSs) from a range of psychiatric, personality, and lifestyle traits to dissect differences in BD sub-phenotypes in two BD cohorts: the Mayo Clinic BD Biobank (N = 968) and Genetic Association Information Network (N = 1001). Participants were assessed for history of psychosis, early-onset BD, rapid cycling (defined as four or more episodes in a year), and suicide attempts using questionnaires and the Structured Clinical Interview for DSM-IV. In a combined sample of 1969 bipolar cases (45.5% male), those with psychosis had higher PRS for SCZ (OR = 1.3 per S.D.; p = 3e-5) but lower PRSs for anhedonia (OR = 0.87; p = 0.003) and BMI (OR = 0.87; p = 0.003). Rapid cycling cases had higher PRS for ADHD (OR = 1.23; p = 7e-5) and MDD (OR = 1.23; p = 4e-5) and lower BD PRS (OR = 0.8; p = 0.004). Cases with a suicide attempt had higher PRS for MDD (OR = 1.26; p = 1e-6) and anhedonia (OR = 1.22; p = 2e-5) as well as lower PRS for educational attainment (OR = 0.87; p = 0.003). The observed novel PRS associations with sub-phenotypes align with clinical observations such as rapid cycling BD patients having a greater lifetime prevalence of ADHD. Our findings confirm that genetic heterogeneity underlies the clinical heterogeneity of BD and consideration of genetic contribution to psychopathologic components of psychiatric disorders may improve genetic prediction of complex psychiatric disorders.

## Introduction

Many psychiatric disorders have moderate to high heritability; however, the genetics of psychiatric disorders are complex and highly polygenic, with each risk variant only conferring a small effect[1]. Psychiatric disorders also have a high level of overlapping clinical heterogeneity, with shared genetic risk explaining some of the clinical overlap, and certain combinations of alleles may contribute to the same psychopathological symptoms in multiple psychiatric disorders. Furthermore, some psychiatric disorders may lie on a continuum rather than being disorders with distinct genetics and biological mechanisms[2, 3].

To accommodate this genetic complexity, investigations of psychiatric disorders have increasingly relied on polygenic risk scores (PRSs), leveraging knowledge from prior large genome-wide association studies (GWASs) to predict genetic risk of particular disorders in a new sample[4]. When the PRS for one disorder is predictive of a second disorder, this indicates a common polygenic contribution to the two disorders[5].

Bipolar disorder (BD) is a complex illness with heterogeneous clinical presentation, and apparent sub-phenotypes often have a different course of illness, prognosis, and treatment response[6–9]. In order to personalize treatment, it is crucial to better understand biological underpinnings of BD clinical sub-phenotypes. One approach is to examine potential relationships of clinical phenotypes to different genetic profiles.

Historically, the relationship between schizophrenia (SCZ) and BD has shaped classification systems in psychiatry[10]. The corresponding link between phenotype and genetics was recently established with the demonstration that BD patients with a history of psychosis, particularly mood incongruent psychosis and psychosis during mania, have increased genetic risk for SCZ[11–16]. However, it is well recognized that BD genetically overlaps– and has high clinical comorbidity with–other major psychiatric conditions, including major depressive disorder (MDD), attention deficit and hyperactivity disorder (ADHD), anxiety disorders, post-traumatic stress disorder (PTSD), obsessive compulsive disorder (OCD), borderline personality disorder and substance use disorders[17–24]. While significant advances have been made in understanding the genetic relationship between BD psychotic sub-phenotypes and SCZ[11–16], little is known about how genetic risks for other psychiatric disorders as well as important personality and lifestyle traits such as body mass index (BMI), risk-taking, and neuroticism relates to psychosis or other BD clinical sub-phenotypes.

The goal of this study was to systematically test if PRSs for major psychiatric conditions and other traits related to BD are predictors of distinct BD sub-phenotypes, in particular with regards to psychosis, age-of-onset, rapid cycling and suicidal behavior. Understanding the shared genetic risk factors between BD clinical sub-phenotypes and other comorbid conditions may contribute to psychiatric clinical classification systems with a more biologically-informed nosological system[25].

## Methods and Materials

### 1. Ethics Statement

Data were collected according to Declaration of Helsinki principles. Mayo Clinic Bipolar Disorder Biobank participants’ consent forms and protocols were reviewed and approved by the Mayo Clinic Institutional Review Board (IRB # 08-008794 00). Opt-in written informed consent was obtained.

### 2. Studies

#### a. Mayo Clinic Bipolar Disorder Biobank

The Mayo Clinic Bipolar Disorder Biobank collection has been described in previous papers[9, 11, 26]. We restricted our analyses to cases with European ancestry (N = 968), because PRSs derived from GWASs of participants with European ancestry perform much worse in non-European ancestries[27]. Sub-phenotypes were determined using the Structured Clinical Interview for DSM-IV (SCID)[28] as well as a patient questionnaire. Detailed information on the assessment of each sub-phenotype can be found in Supplementary Table 1.

**Table 1.** Table of sub-phenotypes and sex for each study. P-value is for a chi-square test of differences between GAIN and Mayo BD-I subject sub-phenotypes.

| Variable | Value | All<br>N = 1969 | GAIN<br>N = 1001 | Mayo<br>N = 968 | p-value |
| --- | --- | --- | --- | --- | --- |
| Sex | Male | 895 (45.5%) | 500 (50.0%) | 395 (40.8%) | 0.003 |
|  | Female | 1074 (54.5%) | 501 (50.0%) | 573 (59.2%) |  |
| BD Type | BD-II | 263 (13.4%) | - | 263 (27.2%) | - |
|  | BD-I | 1706 (86.6%) | 1001 | 705 (72.8%) |  |
| Psychosis | No Psychosis | 880 (47.3%) | 344 (34.9%) | 536 (61.3%) | 3e-7 |
|  | Psychosis | 980 (52.7%) | 642 (65.1%) | 338 (38.7%) |  |
|  | Missing | 109 | 15 | 94 |  |
| Age-of-onset | > 18 yrs | 1292 (70.0%) | 569 (60.3%) | 723 (80.0%) | 1e-13 |
|  | < 19 yrs | 555 (30.1%) | 374 (39.7%) | 181 (20.0%) |  |
|  | Missing | 122 | 58 | 64 |  |
| Rapid Cycling | No | 903 (50.1%) | 508 (60.3%) | 395 (41.1%) | 3e-11 |
|  | Yes | 901 (49.9%) | 334 (39.7%) | 567 (58.9%) |  |
|  | Missing | 165 | 159 | 6 |  |
| Suicide attempts | None | 1115 (57.2%) | 554 (56.5%) | 561 (58.0%) | 0.619 |
|  | 1+ | 833 (42.8%) | 427 (43.5%) | 406 (42.0%) |  |
|  | Missing | 21 | 20 | 1 |  |

#### b. Genetic Association Information Network (GAIN)

The Bipolar Disorder Genome Study Consortium conducted a genome-wide association study (GWAS) of BD as part of the Genetic Association Information Network (GAIN)[29]. We obtained the data from dbGaP (phs000017.v3.p1), and restricted our analyses to cases with European ancestry (N = 1001). All cases met criteria for DSM IV-defined bipolar I disorder (BD-I). Subjects recruited at different times were interviewed with the Diagnostic Interview for Genetic Studies 2, 3, or 4 (DIGS 2, 3, 4). Detailed information on the assessment of each sub-phenotype can be found in Supplementary Table 1.

### 3. Genotyping and Quality Control

#### a. Mayo Clinic Bipolar Biobank

Genotyping and genetic data quality control of this sample was previously described as part of a larger case-control study[11]. Briefly, the Illumina HumanOmniExpress platform was used to genotype 1046 BD cases. For quality control purposes, we excluded subjects with <98% call rate and related subjects. SNPs with call rate <98%, MAF <0.01, and those not in Hardy-Weinberg Equilibrium (HWE; P<1e-06) were removed. After these steps 643 011 SNPs and 968 subjects remained.

#### b. GAIN

Genotyping and quality control procedures for the GAIN-BD data were previously described by Smith et al.[30], Briefly, the Affymetrix Genome-Wide Human SNP Array 6.0 platform was used to genotype cases and after excluding SNPs with call rate <98%, MAF <0.01, and those not in HWE, 726 315 SNPs and 1001 subjects of European ancestry remained.

#### C. Imputation

Genotypes in both the GAIN and Mayo samples were imputed to the 1000 genomes reference panel, as previously described for the GAIN sample[9]. Specifically, SHAPEIT[31] was used for haplotype phasing and imputation was performed using IMPUTE2.2.2[32] with the 1000 genome project reference data (phase 1 data, all populations). Dosage data was converted to best guess genotype for the well-imputed (dosage R^2^ >0.8) and common (MAF >0.01) SNPs, resulting in more than 5 million SNPs in both datasets.

### 4. Polygenic Risk Scores (PRSs)

PRSs were included in the analysis if: 1) there was evidence of significant genetic correlation of the trait with BD and 2) we had at least 80% power to detect PRS association in a general case-only analysis of our data assuming 50% prevalence of the subphenotype. We began by considering PRSs for major psychiatric disorders (BD[33], SCZ[34], MDD[35], ADHD[36], anxiety[37], PTSD[19], OCD[38], anorexia nervosa (AN)[39], alcohol use disorder (AUD)[40], and insomnia[41]) and personality and lifestyle traits related to BD (alcohol consumption[40], educational attainment (EA)[42], risk-taking[43], subjective well-being (SWB)[44], neuroticism[45], anhedonia[46], and body mass index (BMI)[47]). GWAS summary statistics were restricted to well-imputed variants (INFO >0.9) when information on imputation quality was available.

Using LD score regression[48], we estimated the genetic correlation of the above traits with BD[33] (Supplementary Table 2). Insomnia and alcohol consumption didn’t have significant genetic correlation with BD and were therefore excluded from further analysis.

Using the `R` package `avengeme` [49], we estimated that training sample sizes of 20,000 would achieve at least 80% power in our analysis assuming moderate overlap of the trait with the sub-phenotype (genetic covariance = 0.1), high polygenicity (# of independent SNPs = 20,000), and 0.005 α-level to account for multiple testing. The study of OCD included an effective sample size of less than 4000 and was thus excluded from further analysis. The final list of PRSs that were tested for association with BD sub-phenotypes is shown in Supplementary Table 2.

For traits that satisfied our inclusion criteria, the PRS-CS[50] auto setting was applied to estimate SNP weights using a fully Bayesian shrinkage approach that shrinks SNP effects with a continuous shrinkage prior. This setting allows the algorithm to learn the global shrinkage parameter from the data to create one set of weights per PRS and therefore does not require a validation dataset. This setting also reduces the multiple testing of standard PRS analyses that search over many p-value thresholds[51]. PLINK version 1.9[52] was used to create PRSs using the shrunken SNP weights. The PRSs were then standardized to have a mean of zero and standard deviation (SD) of one.

### 5. Statistical Analyses

In each dataset, we performed principal components (PCs) analysis of the genotyped SNPs and kept the first four PCs to be used as within-study nested covariates in subsequent PRS association analyses. In all models, study indicator and an interaction of study and within-study PCs were included as covariates to control for population stratification. All twelve PRSs were individually modeled using a multivariate logistic regression model with each sub-phenotype (psychosis, early-onset BD, rapid cycling, and attempted suicide) as the outcome.

We used 10,000 permutations to find the significance threshold to control the false positive rate testing for association with each sub-phenotype with fourteen PRSs (α = 0.005) as well as the family-wise error rate (α = 0.001). For each sub-phenotype, we also included all significant PRSs (p <0.005) in a joint model, to estimate the relative contribution of the PRSs after adjusting for other important PRSs. We report the variance explained in the sub-phenotype by each PRS after adjustment for other PRSs using Nagelkerke’s pseudo-R^2^ statistic. All statistical analyses were performed in R 3.5.2.

## Results

### Sample Description

Table 1 summarizes the demographic and sub-phenotype information of each study. There was a difference in the sex distribution between the two samples. The GAIN study only included BD type I cases, and the distributions were also significantly different for all sub-phenotypes besides attempted suicide. GAIN BD cases had a higher rate of psychosis and early-onset BD, while Mayo cases had higher rates of rapid cycling, which is more prevalent in women[53].

Figure 1 shows a forest plot of the significant PRS associations with each sub-phenotype further broken down by study. Further detailed results for each sub-phenotype can be found in Supplementary Tables 3-6.

**Figure 1.**
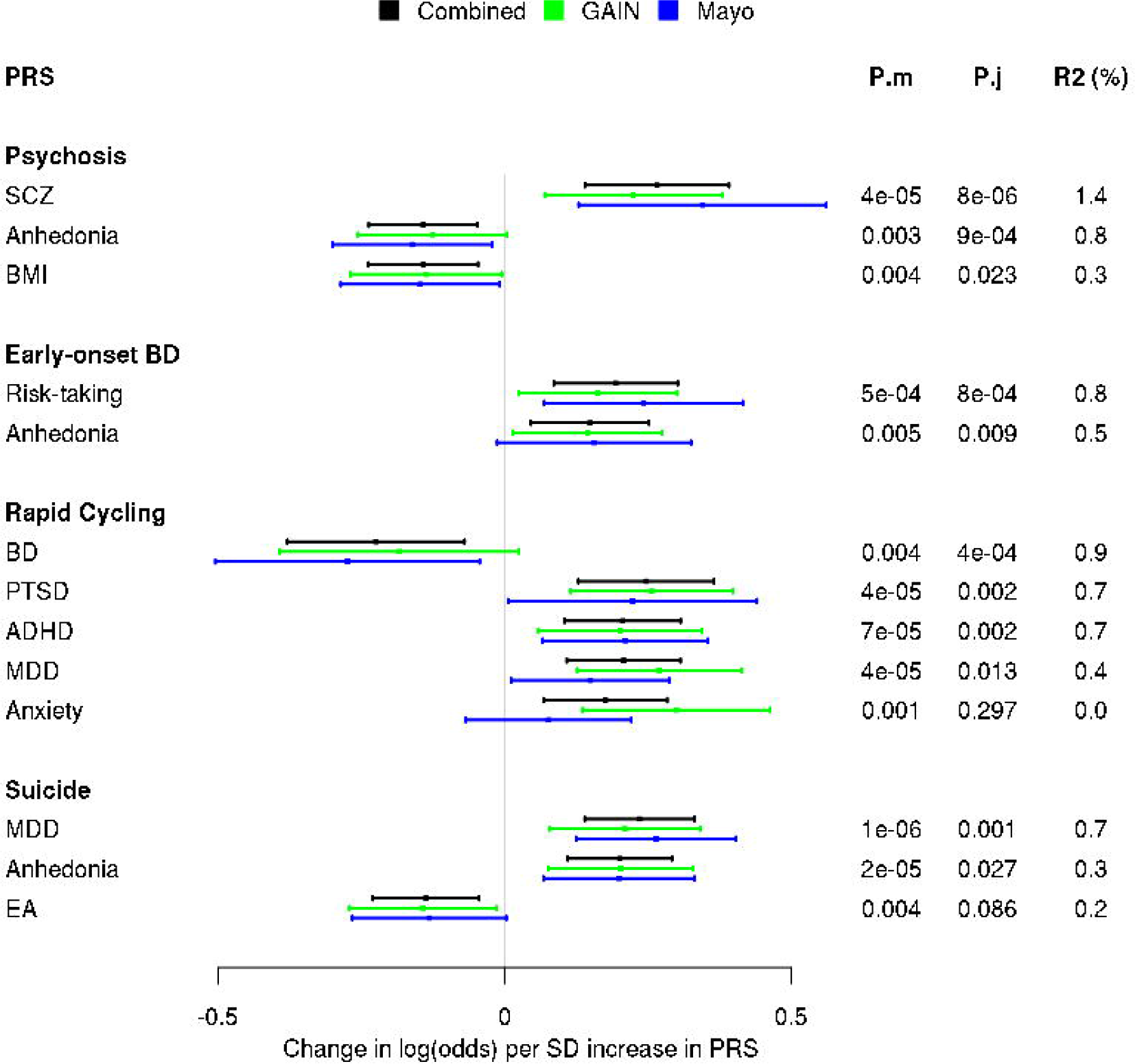
Forest plot of significant PRS associations with each sub-phenotype stratified by study (black = Combined; green = GAIN; maroon = Mayo). Each bar represents a 95% confidence interval of the increased log(odds) in the sub-phenotype associated with one SD increase in the PRS. P-values for each PRS included in the model by itself (P.m) or with other significant PRSs (P.j) and adjusted Nagelkerke’s R^2^ (R2) are listed in the margins for each PRS.

### Psychosis

Cases with psychosis versus no psychosis had higher PRSs for SCZ (OR = 1.3, 95% CI 1.15 to 1.48; p-value = 3.5e-5), but lower PRSs for anhedonia (OR = 0.87, 95% CI 0.79 to 0.95; p-value = 0.003), and BMI (OR = 0.87, 95% CI 0.79 to 0.95; p-value = 0.004). These three PRSs explained 2.6% of the variation in psychosis in the joint model. While anhedonia is a component of MDD and the two PRSs are positively correlated (r = 0.41), the PRS for MDD was not associated with psychosis in BD (OR = 0.96, 95% CI 0.87 to 1.06; p-value = 0.45).

### Early-onset BD

Higher PRSs for risk-taking (OR = 1.21, 95% CI 1.09 to 1.35; p-value = 0.0005; adj. Nagelkerke’s R^2^ = 0.8%) and anhedonia (OR = 1.16, 95% CI 1.05 to 1.29; p-value = 0.0047; adj. Nagelkerke’s R^2^ = 0.8%) were observed in cases with early-onset BD compared to cases that developed BD after age 18.

### Rapid Cycling

Cases with rapid cycling versus those without rapid cycling had higher ADHD PRS (OR = 1.23, 95% CI 1.11 to 1.36; p-value = 7e-5; adj. Nagelkerke’s R^2^ = 0.8%), MDD PRS (OR = 1.23, 95% CI 1.11 to 1.36; p-value = 4e-5; adj. Nagelkerke’s R^2^ = 0.5%), PTSD PRS (OR = 1.28, 95% CI 1.14 to 1.44; p-value = 4e-5; adj. Nagelkerke’s R^2^ = 0.7%), and PRS for anxiety (OR = 1.19, 95% CI 1.07 to 1.33; p-value = 0.001; adj. Nagelkerke’s R^2^ = 0.1%). Cases with rapid cycling also had lower BD PRSs (OR = 0.80, 95% CI 0.68 to 0.93; p-value = 0.004; adj. Nagelkerke’s R^2^ = 0.9%). The five PRSs explained 3.9% of the variation in rapid cycling when included in one model.

### Attempted suicide

The genetic risk for MDD (OR = 1.26, 95% CI 1.15 to 1.39; p-value = 1e-6; adj. Nagelkerke’s R^2^ = 0.7%) and anhedonia (OR = 1.22, 95% CI 1.12 to 1.34; p-value = 2e-5; adj. Nagelkerke’s R^2^ = 0.3%) was higher in cases with at least one suicide attempt versus those with none. Cases with an attempted suicide also had a lower PRS for educational attainment (OR = 0.87, 95% CI 0.79 to 0.96; p-value = 0.0036; adj. Nagelkerke’s R^2^ = 0.2%). The three PRSs explained a total of 2.3% of the variation when included in one model, but only the MDD PRS remained significant after accounting for the other PRS associations.

## Discussion

To our knowledge, this is the first PRS dissection of clinical sub-phenotypes in BD that is comprehensive with respect to the range of psychiatric, personality, and lifestyle phenotypes for which genetic liabilities were estimated and used to predict the BD sub-phenotypes. Previous analyses of BD sub-phenotypes of psychosis, early onset BD and suicide focused on genetic liability to the major psychiatric diagnoses of BD, SCZ, and MDD[6, 11, 12, 16, 54]. Here, we took an expanded agnostic approach to PRS analysis by using many different PRSs beyond just these three to more systematically test for PRS association with clinically important sub-phenotypes of BD, including rapid cycling. Importantly, the contribution of each PRS to a sub-phenotype was assessed after adjusting for the other PRSs’ contributions, thereby assessing how predictive a genetic risk is above and beyond other genetic risks. The forest plots in Figure 1 show that our results were highly comparable in the two cohorts, lending greater confidence to the conclusions. Overall, we find that the different BD clinical sub-phenotypes have different profiles of PRS associations with major psychiatric conditions.

### BD with psychosis

Previously, using the Mayo Clinic sample, we showed that BD patients with a history of psychosis during mania had higher genetic risk for SCZ [11]. Here, this finding is replicated in the GAIN cohort. This finding was also reported by Ruderfer et al.[16], in a larger study that included both the Mayo and GAIN BD cohorts. However, in addition to this relationship, in the present study we also found that BD cases that have not experienced psychotic symptoms had higher genetic scores for anhedonia and BMI. Association of higher genetic risk for anhedonia with a subtype of BD without psychotic features implies that a patient with more genetic predisposition for anhedonia during major depressive episodes is less likely to include episodes with psychotic features. In fact, rates of psychotic features are higher in BD compared with MDD and familial studies show a greater heritability of psychotic features in BD relative to in other mood disorders[55]. Interestingly, we did not observe a significant association with MDD PRS despite a strong genetic correlation between anhedonia and MDD. This may underscore the importance of relying on core symptoms in these analyses, instead of using more complex and syndromal entities like MDD. The relationship between BMI and psychosis is complex and influenced by heritable, environmental, and iatrogenic factors. Over the course of illness, most patients with BD and psychosis gain weight, which contributes to morbidity and mortality[56, 57]. Our finding that BD patients with psychosis have lower genetic predisposition to elevated BMI than BD patients without psychosis suggest that weight gain in those with psychosis likely occurs primarily as a side effect of medications. However, the complex relationship between BD and greater body weight needs to be further explored in the context of sub-phenotypes and use of atypical antispychotics or lithium.

### Early-onset BD

We found evidence that higher genetic liability for risk-taking behavior was associated with early-onset BD, but no evidence that genetic risk for SCZ or BD were associated with age of onset of illness. A previous study of polygenic associations with age-of-onset of BD also showed no association of SCZ or BD genetic risk with both a dichotomous sub-phenotype, as defined in our study, or continuous age-of-onset[58]. Risk-taking is a hallmark feature of normative adolescence, but is also commonly seen in mania. There are several possible explanations for the risk-taking PRS and early-onset BD association found in this study. Perhaps the simplest explanation is that youth with particularly high propensity for risk-taking behaviors come to clinical attention earlier and subsequently have BD identified at an earlier age. However, there are several potential limitations that may have affected these findings. Due to the way the data were collected, for this study, age of onset was dichotomized based on a cutoff age of 18, which may have reduced power. Also, our early-onset BD definition did not differentiate between age of first manic and first depressive episodes. Furthermore, given the high genetic and clinical overlap between BD and other conditions investigated here (e.g. ADHD), a study of age-of-onset of any psychiatric disorder/symptom rather than just BD could be informative. It is of note that earlier onset MDD is associated with more pronounced aggressive/impulsive traits[59]. Nevertheless, the observed association of risk-taking PRS with early vs. late onset BD is intriguing and warrants further investigation.

### BD with rapid cycling

Previous clinical studies have shown a strikingly higher clinical comorbidity rate of ADHD in BD patients with rapid cycling compared to non-rapid cycling BD patients[60]. While the general genetic association between ADHD and BD has been described before[17], our results are the first study to show possible genetic underpinnings for this specific rapid cycling BD and ADHD association. We also found a strong association of MDD genetic risk with rapid cycling. This implies that genetic variation related to ADHD and MDD may also be related to episode frequency in BD, and that comorbid ADHD and more depressive episodes would be clinically associated with the rapid cycling form of BD, though the predominant directionality of mood episodes was not discernible from the available data. Rapid cycling BD has been reported to have more episodes of major depression and a higher rate of parental MDD compared with non-rapid cycling BD[61], which is consistent with our PRS association findings. Finally, rapid cycling cases had lower BD PRS as reported in a previous investigation[15]. This could simply reflect that prevalence of rapid cycling in cases ascertained for the sample used in the GWAS of BD by the Psychiatric Genomics Consortium (PGC) was lower than in the two samples included here, but still demonstrates a systematic difference in genetics of rapid cycling and non-rapid cycling BD.

### BD with a history of a suicide attempt

Our finding of increased MDD genetic load in BD patients with a history of suicide attempts is consistent with a recent study that included both of the Mayo and GAIN data, which showed that genetic risk factors for MDD increase the risk for suicide trans-diagnostically[54]. BD with a history of suicide attempt having a higher MDD genetic liability is consistent with the clinical observation that suicide attempts are most common during major depressive episodes or mixed states and rare during manic episodes or while euthymic[62, 63]. Interestingly, even after adjusting for MDD PRS, we also found that genetic liability for anhedonia is marginally associated with suicide, suggesting that anhedonia may be a particularly relevant factor contributing to suicidality, compared to other components that comprise the MDD syndrome. This is consistent with findings from non-genetic studies, which found that association of anhedonia with suicidality is independent of the association with depression and psychotic features[64].

### Methodological limitations

PRSs used in this study are based on data from previously published large scale investigations and are limited by the diagnostic accuracy, recruitment criteria, and methodology of previous studies. The most recent PGC study of BD[33] included the cases and controls from the GAIN and Mayo samples, and most PGC studies of other disorders used controls from the GAIN study. Sample overlap of testing datasets with training data can create substantial biases in PRS analyses. However, here we studied genetic differences within cases and thus, the sample overlap is not expected to bias our results, because there is no correlation between case-control status used to build the training models and the within-case sub-phenotypes. Another limitation in this study is the lack of data on the number, duration and severity of major depressive and manic episodes to more precisely map the clinical picture onto the PRS profile. Finally, it is important to note that no PRS explained a large amount of variation in our analysis. Thus, while the associations identified in this study provide evidence of genetic differences that may underlie clinical subtypes of BD, these PRSs cannot yet be used for purposes of personalized psychiatry.

## Conclusion

Our findings contribute to the understanding of the underlying genetic causes of clinical heterogeneity of BD and of comorbidity between BD and other major psychiatric conditions. We find evidence that psychopathologic components of BD, including psychotic symptoms, rapid cycling and suicidal behavior are linked to the PRSs for related disorders including schizophrenia, ADHD and MDD, respectively. Finally, larger studies are needed to more precisely map genetic risk factors to clinical sub-phenotypes. Harmonization of sub-phenotypes across studies is a well-recognized challenge. Nevertheless, such efforts are critical in helping to classify psychiatric disorders more accurately and identify risk of suicide, psychosis, and other adverse outcomes in patients.

Supplementary information is available at MP’s website.

## Data Availability

Data is available in our supplemental files.

## Acknowledgments

This work was supported by the Marriott Foundation and the Thomas and Elizabeth Grainger Fund in Bipolar Functional Genomics and Drug Development awarded to the Mayo Clinic. Establishment of the Bipolar Disorder Biobank was supported by a generous gift from the Marriot Family and the Mayo Clinic Center for Individualized Medicine. The contributions from authors JJM, AT, and MMW was supported by NIMH R0MH121921 (Wickramaratne and Mann, M.P.I.s).

## Conflicts of Interest

Dr Frye has received grant support from Assurex Health, Myriad, Pfizer, National Institute of Mental Health (RO1 MH079261), National Institute of Alcohol Abuse and Alcoholism (P20AA017830), Mayo Foundation; has been a consultant to Janssen Global Services, LLC, Mitsubishi Tanabe Pharma Corporation, Myriad, Sunovion, and Teva Pharmaceuticals; has received CME/Travel Support/presentation from CME Outfitters Inc. and Sunovian; Mayo Clinic has a financial interest in AssureRx and OneOme. Dr. McElroy is a consultant to or member of the scientific advisory boards of Bracket, MedAvante, Naurex, Shire, and Sunovion. She is a principal or co-investigator on studies sponsored by the Agency for Healthcare Research & Quality (AHRQ), AstraZeneca, Cephalon, Forest, Marriott Foundation, National Institute of Mental Health, Orexigen Therapeutics, Inc., Shire, and Takeda Pharmaceutical Company Ltd. She is also an inventor on United States Patent No. 6,323,236 B2, Use of Sulfamate Derivatives for Treating Impulse Control Disorders, and along with the patent’s assignee, University of Cincinnati, Cincinnati, Ohio, has received payments from Johnson & Johnson, which has exclusive rights under the patent. In the last three years, Dr Weissman has received research funds from NIMH, Templeton Foundation, Brain and Behavior and the Sackler Foundation and has received royalties for publications of books on interpersonal psychotherapy from Perseus Press, Oxford University Press, on other topics from the American Psychiatric Association Press and royalties on the social adjustment scale from Multihealth Systems. None of these represent a conflict of interest.

